# Decline of Antimicrobial Resistance in *Pseudomonas aeruginosa* Bacteremia Following the COVID-19 Pandemic: a Longitudinal Observational Study

**DOI:** 10.1101/2025.01.11.25320374

**Authors:** Yulia Butscheid, Pascal M. Frey, Marc Pfister, Lisa Pagani, Roger D. Kouyos, Thomas C. Scheier, Willy I. Staiger, Stefano Mancini, Silvio D. Brugger

**Affiliations:** Department of Infectious Diseases and Hospital Epidemiology, University Hospital Zurich, University of Zurich, Zurich, Switzerland; Department of General Internal Medicine, Bern University Hospital, Inselspital, University of Bern, Bern, Switzerland; Institute of Integrative Biology, ETH Zurich, Zurich, Switzerland; Institute of Medical Microbiology, University of Zurich, Zurich, Switzerland

**Keywords:** *Pseudomonas aeruginosa*, bloodstream infection, multidrug resistance, COVID-19

## Abstract

**Background:** Multidrug-resistant Pseudomonas aeruginosa (MDRPA) poses significant challenges in hospital settings. Understanding the effects of the unprecedented changes brought by the COVID-19 pandemic on antimicrobial resistance in P. aeruginosa (PA) is essential to inform stewardship efforts. This study investigated the dynamics of antimicrobial resistance in PA bacteremia after the start of the pandemic.

**Methods:** This single-centre retrospective cohort study included adult inpatients with PA bacteraemia at the University Hospital Zurich between January 2014 and December 2023. Data were retrieved from electronic records. The primary outcome was the association between the start of the COVID-19 pandemic and PA with MDR, defined as resistance to ≥3 of 5 antibiotic classes. We used logistic regression to adjust for age, sex and ICU treatment, accounting for multiple bacteremia instances within the same patient using cluster-robust standard errors. Secondary outcomes included changes in resistance patterns and patient demographics, with antimicrobial exposure assessed as median monthly days of therapy (DOT).

**Results:** A total of 493 instances of PA bacteremia in 333 patients were observed during the study period. The proportion of MDRPA declined from 21% (62/291) pre-pandemic to 9% (19/202) post-pandemic (adjusted OR 0.38, 95% CI 0.18–0.79, p=0.01). The occurrence of MDRPA during hospitalisation following an initial instance of non-MDRPA bacteremia was rare and unlikely to happen earlier than after two weeks. Antimicrobial consumption patterns shifted after the start of the pandemic, with reduced use of amikacin and ciprofloxacin and increased use of cefepime and meropenem. Overall inhospital-mortality among patients with MDRPA bacteraemia remained high (28%), with no substantial differences before and after the pandemic (adjusted hazard ratio 1.57, 95% CI 0.43–5.67, p=0.49).

**Conclusion:** We observed a decline in MDRPA occurrence after the start of the COVID-19 pandemic, possibly driven by intensified infection control measures, shifts in antimicrobial use, and changes in patient populations.

## INTRODUCTION

Multidrug-resistant *Pseudomonas aeruginosa* (MDRPA) is a major public health challenge in hospital settings, where it contributes significantly to morbidity, mortality, and healthcare costs [1,2]. *P. aeruginosa* is an opportunistic pathogen responsible for a wide range of healthcare-associated infections, including pneumonia, urinary tract infections, surgical site or wound infections, and bacteraemia. The latter, in particular, represents a severe manifestation and is associated with high mortality rates of >30% in critically ill patients [3,4].

Designated as a high priority pathogen by the World Health Organization [5], MDRPA requires focused efforts to mitigate its impact [6]. Effective strategies include enhanced infection prevention and control measures, such as rigorous hand hygiene protocols, environmental cleaning, and antimicrobial stewardship.

The COVID-19 pandemic introduced unprecedented changes in hospital settings to limit the spread of SARS-CoV-2 that included stricter infection control protocols, shifts in patient demographics and population behaviour, and altered patterns of antimicrobial consumption, all of which likely influenced the dynamics of antimicrobial resistance [7]. Nevertheless, concerns have been raised that the pandemic may have accelerated the threat of antimicrobial resistance, primarily due to widespread antibiotic overuse [8,9]. Increased antibiotic prescribing in hospitals, particularly for pneumonia, has been associated with a rise in multidrug-resistant gram-negative organisms, illustrating the potential role of the pandemic in accelerating antimicrobial resistance [10].

Although the direction of antimicrobial resistance (AMR) trends during the COVID-19 pandemic had been hypothesised to mainly increase, uncertainty remains regarding actual AMR measures, with opposing forces potentially influencing outcomes [7]. Enhanced infection control measures and reduced patient mobility were hypothesised to limit the spread of resistant pathogens [11], while increased antibiotic use was expected to sustain selective pressure, potentially worsening resistance. Another potential factor influencing AMR in *P. aeruginosa* during the COVID-19 pandemic is the shift in patient demographics, particularly when considering the high mortality rates observed in individuals with chronic pulmonary disease. This may have led to a reduction in the population prone to recurrent *P. aeruginosa* infections and repeated exposure to antimicrobial treatments, potentially altering the selective pressures driving resistance. These contrasting influences underscore the equipoise surrounding AMR trends during the pandemic, particularly for MDRPA, a key nosocomial pathogen with a high capacity for adaptation to selective pressures.

We aimed to investigate the association between pandemic-related changes and MDRPA occurrence and characterise dynamics in resistance patterns and patient demographics before and after the start of the COVID-19 pandemic. Based on previous literature suggesting a potential increase of AMR during the pandemic [8,10], we hypothesised an increased occurrence of MDR in *P. aeruginosa* bacteremia after the start of the pandemic.

## METHODS

### Study Design and Population

This single-centre retrospective cohort study included all inpatients aged ≥18 years with at least one blood culture positive for *P. aeruginosa* (PA) obtained for microbiological testing at the University Hospital Zurich between January 2014 and December 2023. Patients were required to have at least one instance (defined as a day with at least one positive blood culture sample) of PA bacteraemia to be included in the cohort. For the purposes of this study, an "instance" of PA bacteraemia was defined as any day with a positive blood culture for PA, irrespective of the number of positive cultures obtained on that day. Subsequent positive cultures occurring on different days were considered separate instances. Due to the lack of information required to differentiate between new infections and persistent bacteraemia, we did not define a minimum interval of days between separate instances of bacteraemia in a pragmatic approach. Outpatients were excluded.

### Data Collection

Clinical data were collected through the use of an in-hospital electronic medical records database (KISIM Version 5.5, Cistec AG, Zurich, Switzerland). Microbiological samples were collected by healthcare workers as ordered by the treating physicians. Samples were processed at the Institute for Medical Microbiology of the University of Zurich. Standard clinical microbiology analytic techniques were used for culturing, isolation and identification of microorganisms as previously described [12]. Data on antimicrobial use were obtained from the hospital’s internal data warehouse, which stores detailed records of individual prescriptions and administrations for each patient. This patient-level data has been systematically recorded since 2018, enabling precise tracking of antimicrobial consumption.

### Primary Outcome

The primary outcome of this study was the association between the time period in relation to the start of the COVID-19 pandemic and the occurrence of MDRPA thereafter in any instance of PA bacteraemia. MDR was defined as resistance to ≥3 out of the following 5 antibiotic classes: i) piperacillin/tazobactam (acylaminopenicillins); ii) ceftazidime and cefepime (cephalosporins); iii) ciprofloxacin and levofloxacin (fluoroquinolones); iv) ≥2 out of amikacin, gentamicin, or tobramycin (aminoglycosides); and v) imipenem or meropenem (carbapenems).

In March 2024 the fourth criterion of the MDR definition for PA was changed to iv) amikacin or tobramycin (gentamicin was excluded based on EUCAST revision). Although this did not directly affect our study period, we nevertheless added a sensitivity analysis to investigate whether our MDR classification was robust regarding this change in MDR definition.

### Other Outcomes

Secondary outcomes included the characterisation of patients with or without MDRPA bacteraemia, focusing on first PA bacteraemia instances only, to compare demographics and clinical characteristics. To describe changes in the characteristics of patients with MDRPA before and after the onset of the COVID-19 pandemic, we further analysed patients who experienced a first MDRPA bacteraemia regardless of any earlier non-MDRPA instances. Additional analyses included in-hospital mortality, changes in resistance to individual antimicrobial agents, as well as changes in antibiotic consumption before and after the start of the COVID-19 pandemic.

### Time period before and after the start of the COVID-19 pandemic

The study period was divided into two distinct time periods: before the start of the COVID-19 pandemic (January 2014–December 2019) and after it (January 2020–December 2023). A precise definition of the pandemic’s duration was avoided, as it was not expected to substantially improve the precision of the study’s findings but would have made yearly comparisons and interpretations more challenging. The start of the pandemic was chosen as the cutoff due to significant changes in hospital operations associated with the pandemic, including enhanced infection control measures, shifts in patient demographics, and increased awareness of antimicrobial stewardship. Additionally, the onset of the pandemic may have led to altered antimicrobial consumption patterns, potentially influencing changes in multidrug resistance.

### Antimicrobial consumption

Antimicrobial consumption was measured as the median number of days of therapy (DOT) per month for each antibiotic, representing the median number of days per month patients were administered a specific antibiotic. This metric was chosen because it provides a more accurate measure of bacterial exposure to antibiotics compared to other approaches better used for dispensing data such as defined daily doses [13].

### Statistical Analysis

Categorical variables were compared using chi-squared tests or Fisher’s exact tests, as appropriate, while continuous variables were analysed using Wilcoxon rank-sum tests. The associations between the time period in relation to the start of the COVID-19 pandemic and the occurrence of MDRPA as well as single antimicrobial resistances in any instance of PA bacteraemia were investigated using logistic regression models, adjusted for patient age, sex, and ICU admission. To account for the non-independence of multiple instances per patient, cluster-robust standard errors were used. This approach was chosen over a mixed-effects model to provide a measure of effect (odds ratios) that is more intuitive and convenient for interpretation.

The Kaplan-Meier method was used to estimate the time to in-hospital development of MDRPA bacteraemia following a first non-MDRPA bacteraemia during the same hospitalisation, as well as in-hospital mortality among patients with first MDRPA bacteraemia. A Cox proportional hazards model was applied to evaluate the association between the pandemic period and in-hospital mortality, adjusting for age, sex, and ICU admission.

A sensitivity analysis was performed by applying both the original and updated MDR definitions to all data to evaluate whether the change in classification criteria affected the identification of MDRPA cases or the study outcomes. In case of missing data for resistance measures in all substances of an MDR-defining class, a further sensitivity analysis of best and worst case scenarios (all missings sensitive or resistant) was planned.

A two-tailed p-value of <0.05 was considered statistically significant without any adjustments for multiple hypothesis testing due to the exploratory nature of this study.

## RESULTS

### Cohort characteristics and MDR in patients with PA bacteremia

A total of 333 adult patients with a first PA bacteraemia between January 2014 and December 2023 were included in our cohort, contributing a total of 493 instances of any PA bacteremia. The median age of the patients was 62 years (interquartile range [IQR] 51–73 years), and 109 (33%) of the included patients were female. Eighty-two (25%) of first PA bacteraemia instances were detected in the intensive care unit (ICU). MDRPA was detected in 39 (12%) of the 333 first PA bacteraemia instances. Patients with MDRPA were younger than those with non-MDRPA (median age 54 vs 64 years), but with a similar distribution of sex. The duration of the hospital stay (26 vs 15 days) as well as the proportion of ICU admissions were significantly higher (51% vs 21%) in patients with MDRPA compared to those with non-MDRPA (**Table 1**).

**Table 1:**
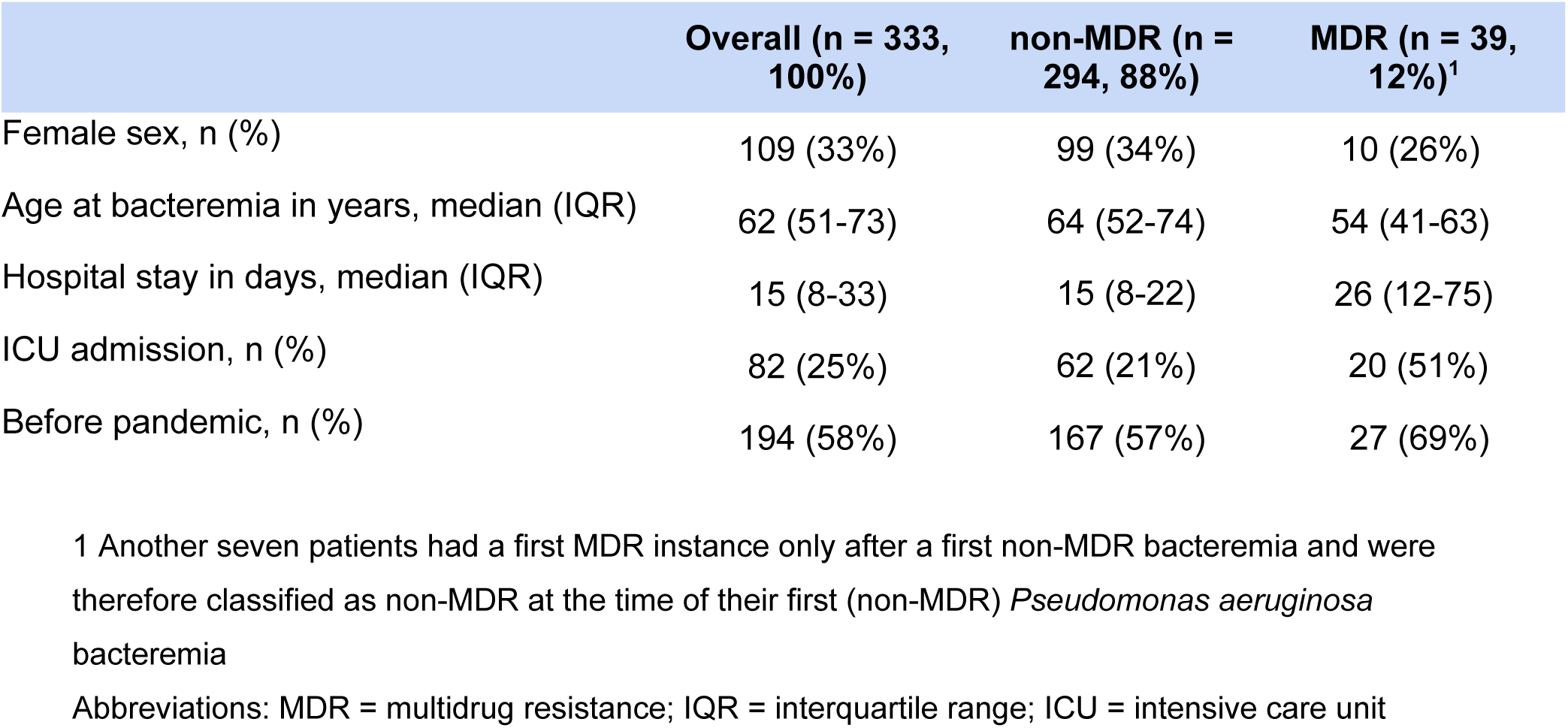
Characteristics of patients with any first PA bacteremia.

|  | Overall (n = 333,<br>100%) | non-MDR (n =<br>294, 88%) | MDR (n = 39,<br>12%)<sup>1</sup> |
| --- | --- | --- | --- |
| Female sex, n (%) | 109 (33%) | 99 (34%) | 10 (26%) |
| Age at bacteremia in years, median (IQR) | 62 (51-73) | 64 (52-74) | 54 (41-63) |
| Hospital stay in days, median (IQR) | 15 (8-33) | 15 (8-22) | 26 (12-75) |
| ICU admission, n (%) | 82 (25%) | 62 (21%) | 20 (51%) |
| Before pandemic, n (%) | 194 (58%) | 167 (57%) | 27 (69%) |

1 Another seven patients had a first MDR instance only after a first non-MDR bacteremia and were therefore classified as non-MDR at the time of their first (non-MDR) *Pseudomonas aeruginosa* bacteremia
Abbreviations: MDR = multidrug resistance; IQR = interquartile range; ICU = intensive care unit

In addition to 39 patients with MDRPA bacteremia as their first PA bacteremia, another 7 patients experienced a subsequent bacteremia instance with an MDRPA after a first non-MDRPA, resulting in overall 46 patients with a first MDRPA bacteremia within the 10-year period. The median age of all patients at their first MDRPA bacteremia was 53 years (IQR 41–62 years) and 13 (28%) patients were female. The median hospital stay of patients with MDRPA bacteremia was 33 days (IQR 12–75 days) and 24 (52%) patients were admitted to the ICU anytime during the hospitalisation. The most common main diagnosis of patients with MDRPA bacteremia was a burn injury, accounting for 15 (33%) patients, followed by malignancy and a solid organ transplantation with 11 (24%) and 8 (17%) patients, respectively. Resistance against 3, 4 and 5 antibiotic classes was detected in 19 (41%), 16 (35%), and 11 (24%) patients, respectively (**Table 2**).

**Table 2:** Characteristics of patients with MDRPA bacteremia.

|  | **Overall**<br>**N=46**<br>**(100%)** | **Before**<br>**pandemic**<br>**n=33 (72%)** | **After start of**<br>**pandemic**<br>**n=13 (28%)** |
| --- | --- | --- | --- |
| Age at admission in years, median (IQR) | 53 (34-62) | 50 (34-60) | 59 (48-65) |
| Female sex, n (%) | 13 (28) | 10 (30) | 3 (23) |
| BMI, median (IQR) | 25 (20-27) | 25 (20-28) | 23 (21-25) |
| Hospital stay in days, median (IQR) | 33 (12-80) | 33 (12-75) | 35 (14-80) |
| ICU admission, n (%) | 24 (52) | 20 (61) | 4 (31) |
| Previous non-MDR bacteremia, n (%) | 7 (15) | 6 (18) | 1 (8) |
| Main Diagnosis: |  |  |  |
| Burn injury, n (%) | 15 (33) | 13 (39) | 2 (15) |
| Organ transplant<sup>1</sup>, n (%) | 8 (17) | 7 (21) | 1 (8) |
| Malignancy, n (%) | 11 (24) | 5 (15) | 6 (46) |
| Primary infection, n (%) | 7 (15) | 3 (9) | 4 (31) |
| Cardo-vascular disease, n (%) | 5 (11) | 5 (15) | 0 (0) |
| Comorbidities (multiple per patient): |  |  |  |
| Arterial hypertension, n (%) | 16 (35) | 9 (27) | 7 (54) |
| Diabetes mellitus, n (%) | 12 (26) | 10 (30) | 2 (15) |
| Chronic kidney disease, n (%) | 9 (20) | 5 (15) | 4 (31) |
| Liver cirrhosis, n (%) | 1 (2) | 1 (3) | 0 (0) |
| Cardo-vascular disease, n (%) | 18 (39) | 11 (33) | 7 (54) |
| Pulmonary disease, n (%) | 8 (17) | 8 (24) | 0 (0) |
| Endocrinopathy, n (%) | 7 (15) | 3 (9) | 4 (31) |
| Chronic infection, n (%) | 8 (17) | 5 (15) | 3 (23) |
| Gastrointestinal disease, n (%) | 1 (2) | 0 (0) | 1 (8) |
| Psychiatric disorder, n (%) | 10 (22) | 8 (24) | 2 (15) |
| Immunosuppression, n (%) | 16 (35) | 11 (33) | 5 (38) |
| In-hospital MDR acquisition, n (%) | 6 (13) | 5 (15) | 1 (8) |
| Resistance to MDR antimicrobial classes: |  |  |  |
| 3 classes | 19 (41) | 13 (39) | 6 (46) |
| 4 classes | 16 (35) | 11 (33) | 5 (38) |
| 5 classes | 11 (24) | 9 (27) | 2 (15) |

1. Including 5 cystic fibrosis patients with lung transplant.
Abbreviations: IQR = interquartile range; BMI = body mass index; ICU = intensive care unit; MDR = multidrug-resistant

Development of an in-hospital MDRPA bacteremia during the same hospital stay after the first non-MDR bacteremia was only observed in 6 patients and seems unlikely to appear in the first two weeks of hospitalisation (**Figure 1**).

**Figure 1:**
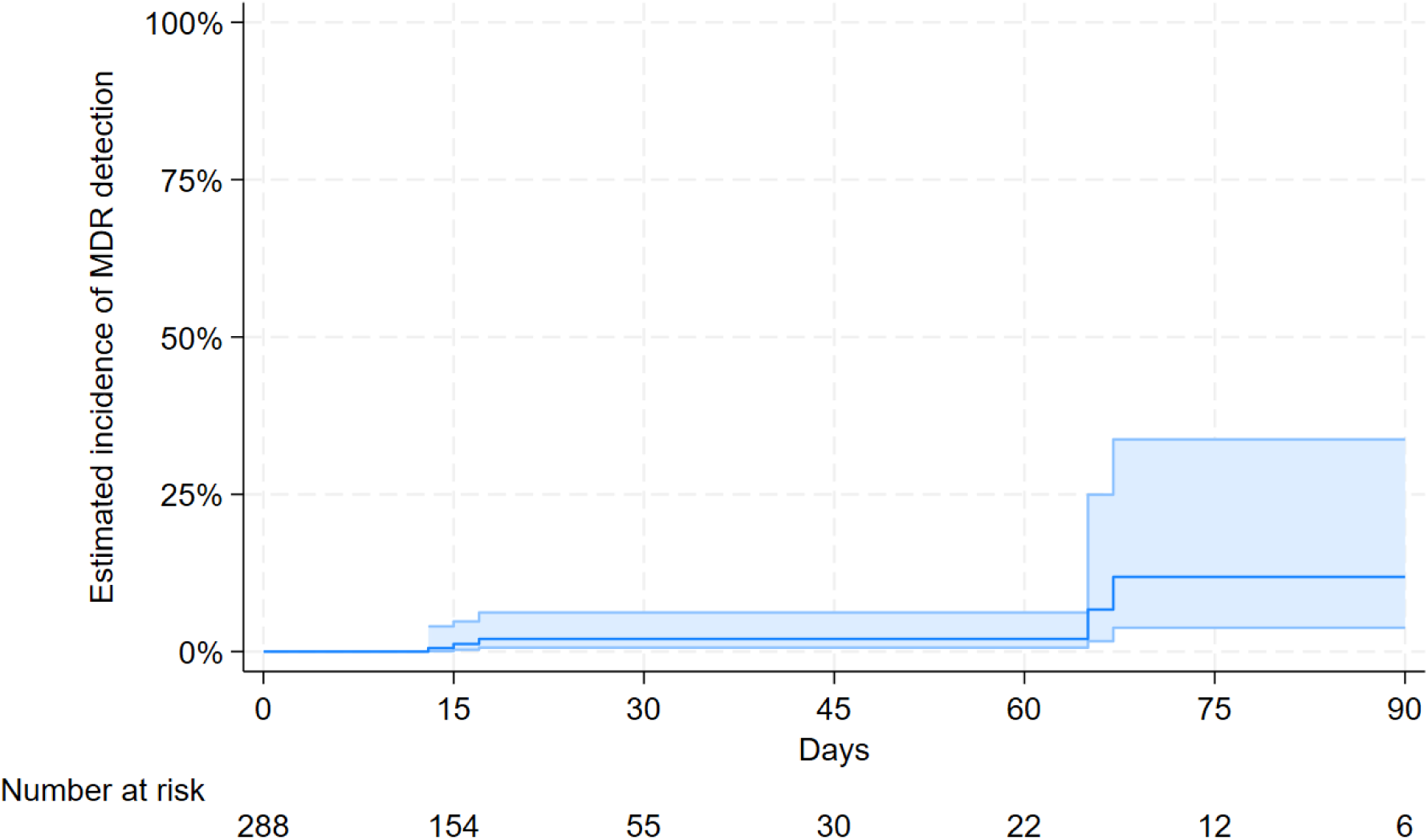
Kaplan-Meier estimates of MDRPA bacteremia incidence during hospital stay after the first non-MDRPA bacteremia. The cumulative incidence curve illustrates the time to in-hospital development of MDRPA bacteraemia during the same hospitalisation after a first non-MDRPA bacteraemia. The absence of events occurring within the first two weeks of hospitalisation suggest that in-hospital acquisition of MDRPA is unlikely during this early period. Time is displayed in days since the initial non-MDRPA bacteraemia instance.

Excluding gentamicin from the MDR definition did not result in any changes of MDR classification.

### Characteristics in patients with PA bacteremia and multidrug resistance before and after the start of the COVID-19 pandemic

Out of 333 patients, 194 (58%) experienced their first PA bacteremia instance before the start of the COVID-19 pandemic and 139 (42%) patients thereafter.

Of the total 46 patients who experienced a first MDRPA bacteremia at any time point between 2014 and 2023, including subsequent events after their first non-MDRPA bacteremia instance, only 13 (28%) patients with MDRPA bacteremia were observed after the start of the pandemic.

Overall, during the investigated period the total annual number of patients with a first PA bacteremia remained stable at around 30-40 cases per year. However, in the years following the start of the COVID-19 pandemic a substantial decline in the proportion of a MDRPA in a first PA bacteremia was observed: from 14% to 9% during the years after 2020, with only 3% in the year 2023 (**Figure 2**).

**Figure 2.**
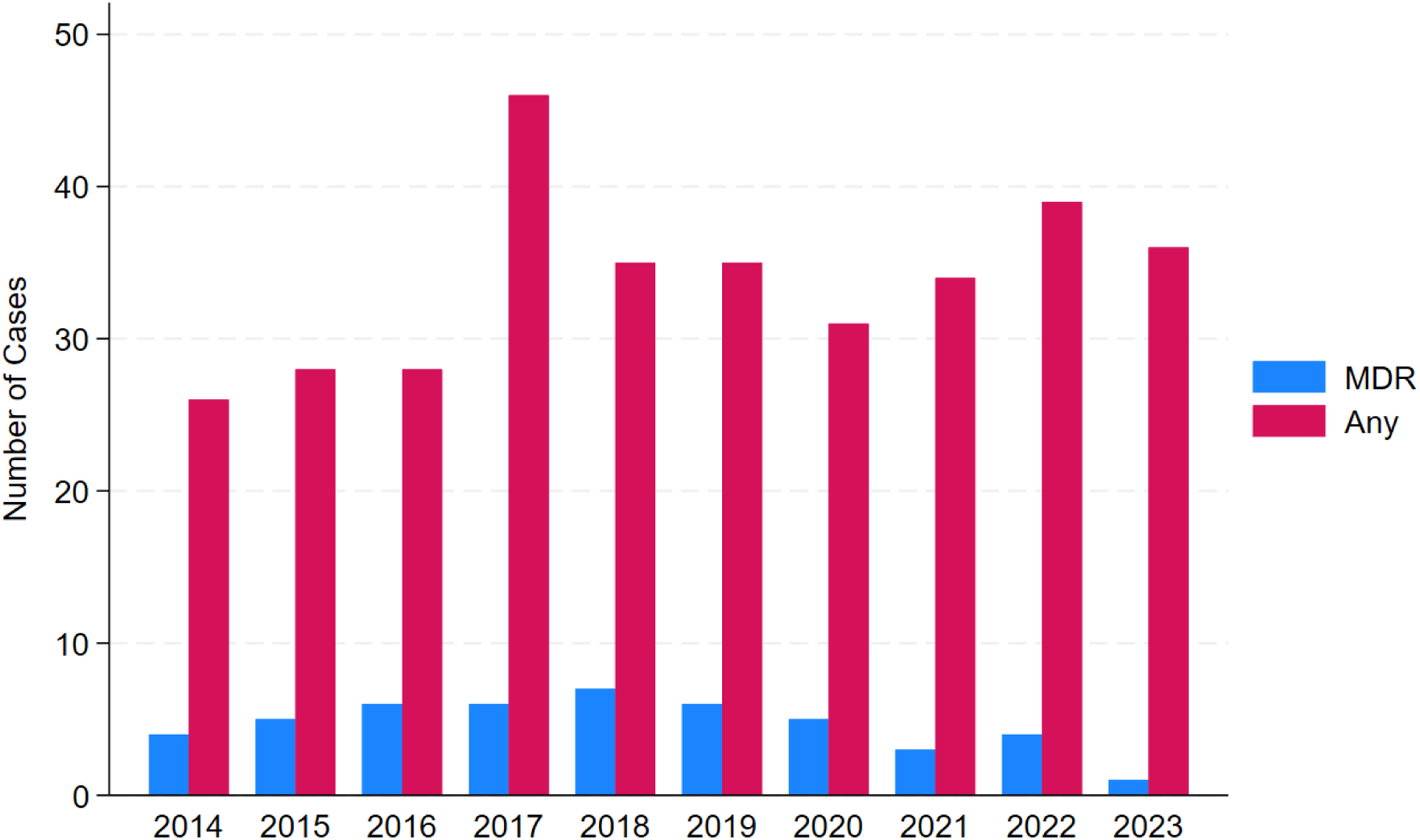
Panel A: Number of first PA bacteremia cases stratified by year. Total annual number of patients with first PA bacteraemia instance from 2014 to 2023, showing stable overall PA occurrence despite changes in MDRPA proportions over the study period.

**Figure 2.**
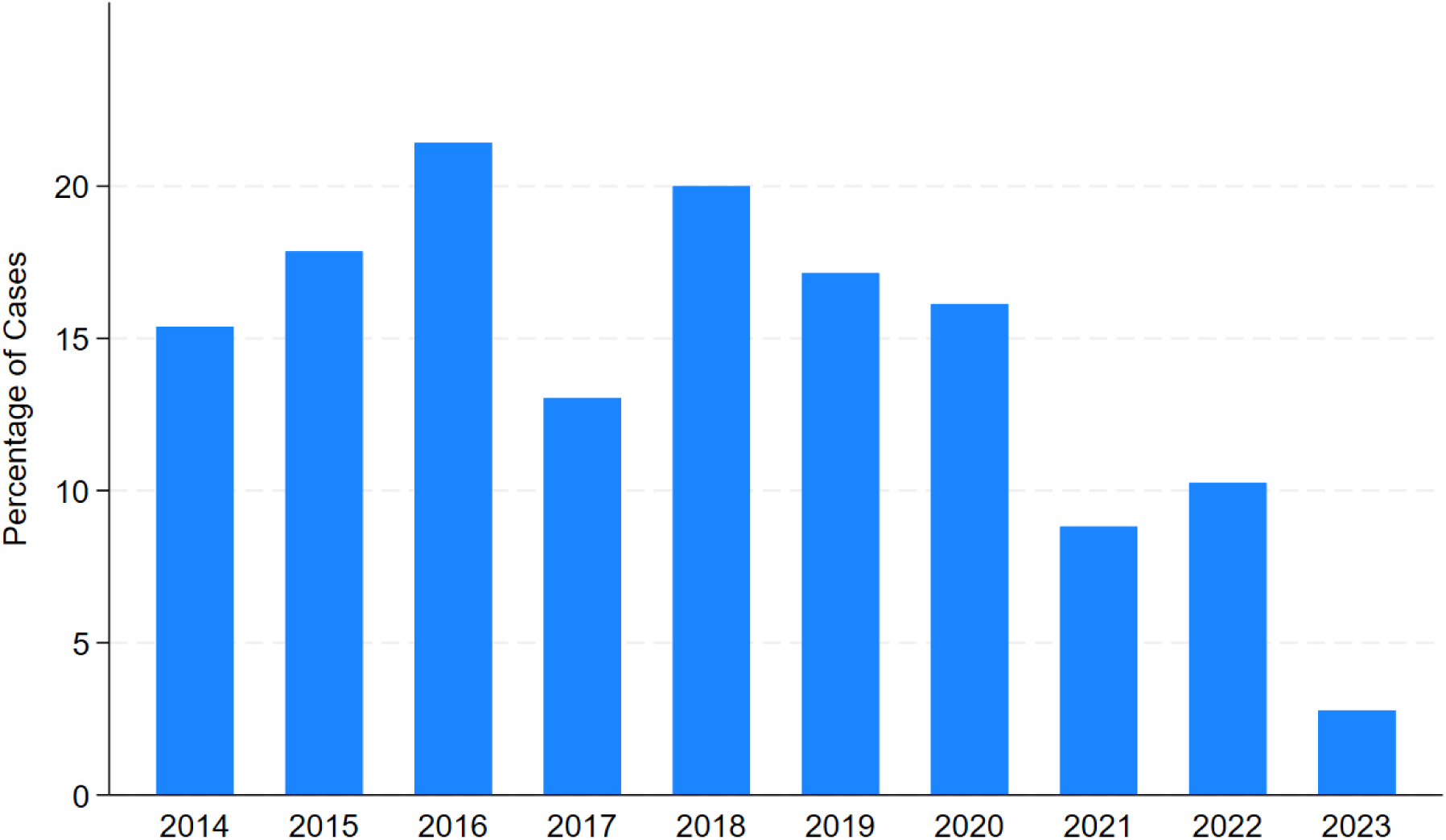
Panel B: Percentage of MDRPA in first bacteremia cases stratified by year. Annual proportion of multidrug-resistant MDRPA among first PA bacteraemia instances from 2014 to 2023. A substantial decline in MDRPA proportions is observed after the start of the pandemic in 2020, with only 3% detected in 2023.

The demographic characteristics of patients with MDRPA bacteremia showed some relevant differences after the start of the COVID-19 pandemic without any patients with main diagnosis of a cardio-vascilar disease as well as pulmonary comorbidities, and substantially less patients with a main diagnosis of either burn injury or organ transplant (**Table 2**).

In-hospital mortality among patients with a first MDRPA bacteraemia was 28% (13/46) in the cohort. Mortality rates were 24% (8/33) in the pre-pandemic period and 38% (5/13) after the start of the pandemic. After adjusting for age, sex, and ICU admission, no significant difference in in-hospital mortality was observed between the periods (HR 1.57, 95%CI 0.43–5.67, p=0.49; **Figure 3**).

**Figure 3:**
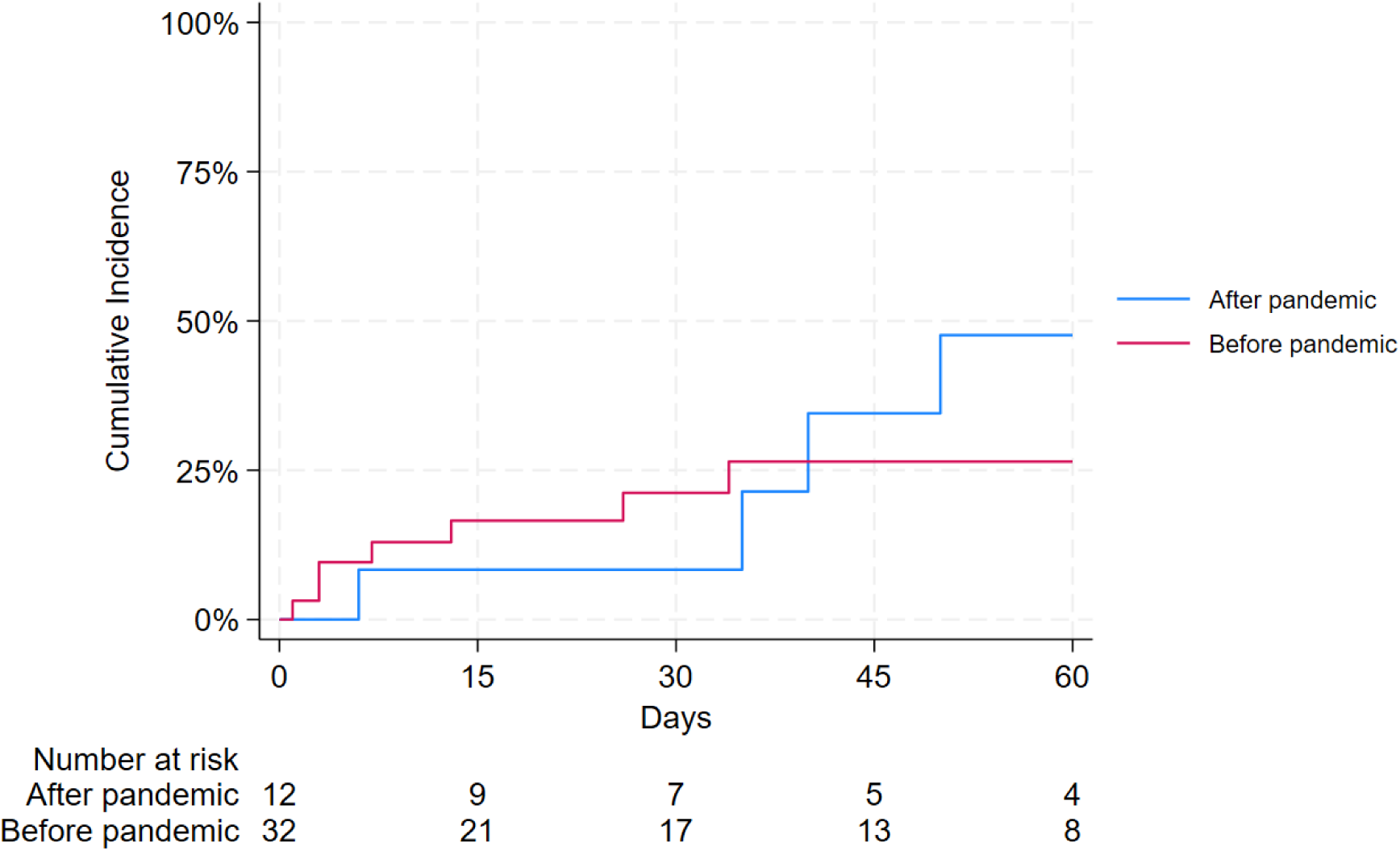
Kaplan-Meier estimates of in-hospital death in patients with a first MDRPA bacteremia before and after start of the pandemic. The Kaplan-Meier curve shows the cumulative probability of in-hospital mortality for patients with first MDRPA bacteraemia, stratified by the pre-pandemic (2014–2019) and pandemic/post-pandemic (2020–2023) periods. Mortality estimates are displayed over time, with no substantial differences in mortality observed between the two periods (HR 1.57, 95%CI 0.43-5.67, p=0.49; adjusted for sex, age and intensive care unit treatment).

### Start of the COVID-19 pandemic and antimicrobial resistance over all instances of PA bacteremia beyond the first per patient

Within the previously described 333 patients, we observed 81 instances (separate days) of MDRPA bacteremia in the total of 493 bacteremia instances over the study period. The proportion of MDR in any of these PA bacteremia instances (including those after the first one in the same patient) declined from 21% MDR before COVID to 9% MDR after the start of the pandemic. Similarly, resistance to individual antimicrobial agents showed varying proportions before and after the pandemic: resistance to amikacin decreased from 11% to 1%, cefepime from 21% to 15%, ceftazidime from 26% to 18%, and ciprofloxacin from 22% to 18% (**Table 3**). We found evidence of an association between the start of the COVID-19 pandemic and the occurrence of MDRPA: the odds of MDR were substantially lower after the start of the pandemic (odds ratio [OR] 0.38, 95%CI 0.18–0.79, p=0.01), adjusting for age, sex, and ICU treatment while taking the non-independence of multiple instances per patient into account (**Table 3** and **Figure 4**). The results from our unadjusted analysis were very similar (OR 0.38, 95%CI 0.19–0.78, p = 0.009).

**Figure 4:**
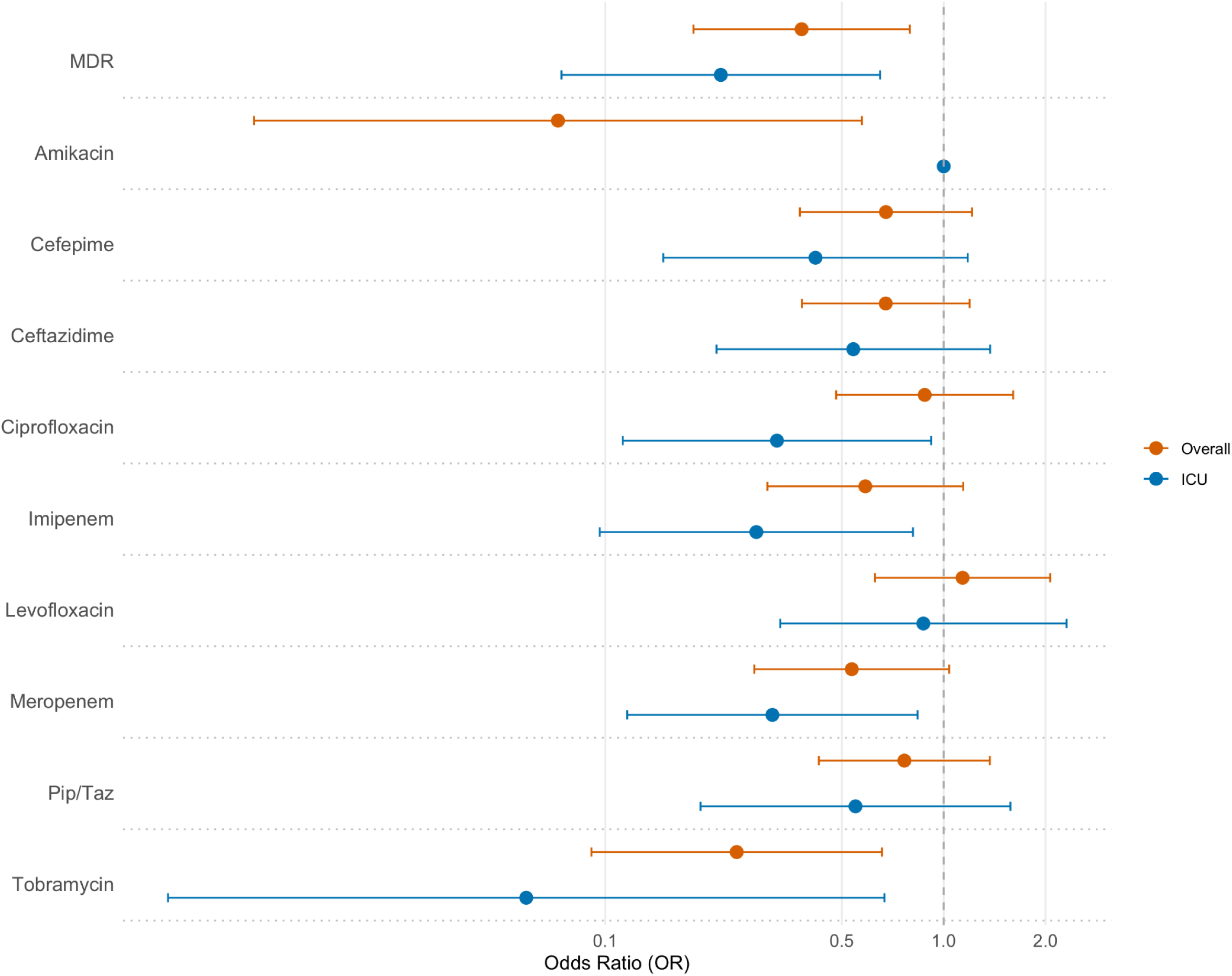
Antimicrobial resistance before and after the start of the COVID-19 pandemic. Forest plot of odds ratios (OR) and 95% confidence intervals (CI) adjusted for intensive care unit (ICU) treatment, sex and age (per 5 years) and taking clustering of multiple instances of bacteremia per patient into account. ORs for the ICU population were not adjusted for ICU treatment.

**Table 3:** Antimicrobial resistance before and after the start of the pandemic.

| Antimicrobial substance<sup>1</sup> | All samples (N = 493) | Before pandemic<sup>2</sup> (n = 291) | After start of pandemic<sup>2</sup> (n = 202) | Adjusted<sup>3</sup> OR (95% CI) | Adjusted<sup>3</sup> p-value |
| --- | --- | --- | --- | --- | --- |
| **MDR** | 81 (16%) | 62 (21%) | 19 (9%) | 0.38 (0.18-0.79) | 0.01 |
| only on ICU | 50 (34%) | 43 (41%) | 7 (17%) | 0.22 (0.07-0.65) | <0.01 |
| **Cefepime** | 92 (19%) | 62 (21%) | 30 (15%) | 0.68 (0.38-1.21) | 0.16 |
| only on ICU | 43 (29%) | 35 (33%) | 8 (19%) | 0.42 (0.15-1.18) | 0.1 |
| **Ceftazidime** | 110 (22%) | 74 (26%) | 36 (18%) | 0.67 (0.38-1.17) | 0.15 |
| only on ICU | 50 (34%) | 39 (37%) | 11 (26%) | 0.54 (0.21-1.37) | 0.2 |
| **Piperacillin / Tazobactam** | 105 (21%) | 68 (23%) | 37 (18%) | 0.77 (0.43-1.36) | 0.3 |
| only on ICU | 46 (31%) | 36 (34%) | 10 (24%) | 0.55 (0.19-1.58) | 0.3 |
| **Imipenem** | 119 (24%) | 83 (29%) | 36 (18%) | 0.59 (0.30-1.14) | 0.12 |
| only on ICU | 69 (47%) | 57 (54%) | 12 (29%) | 0.28 (0.10-0.81) | 0.02 |
| **Meropenem** | 105 (21%) | 76 (26%) | 29 (14%) | 0.53 (0.27-1.03) | 0.06 |
| only on ICU | 67 (46%) | 55 (52%) | 12 (29%) | 0.31 (0.12-0.84) | 0.02 |
| **Ciprofloxacin** | 100 (20%) | 63 (22%) | 37 (18%) | 0.88 (0.48-1.60) | 0.7 |
| only on ICU | 53 (36%) | 44 (42%) | 9 (21%) | 0.32 (0.11-0.92) | 0.034 |
| **Levofloxacin** | 110 (22%) | 65 (22%) | 45 (22%) | 1.14 (0.62-2.10) | 0.7 |
| only on ICU | 58 (39%) | 42 (40%) | 16 (38%) | 0.87 (0.33-2.31) | 0.8 |
| **Amikacin** | 14 (6%) | 13 (11%) | 1 (1%) | 0.07 (0.01-0.57) | 0.01 |
| only on ICU | 9 (6%) | 9 (9%) | 0 (0%) | N/A | N/A |
| **Tobramycin** | 41 (8%) | 35 (12%) | 6 (3%) | 0.24 (0.09-0.66) | < 0.01 |
| only on ICU | 27 (18%) | 26 (25%) | 1 (2%) | 0.06 (0.01-0.67) | 0.02 |

Each separate day with at least one positive blood culture was considered to be an instance of bacteremia. Analyses with confidence intervals and p-values have taken non-independence of repeat instances per patient into account.
<sup>1</sup> Missing resistance testing results for: ceftazidime in 1, tobramycin in 1, levofloxacin in 2 and amikacin in 247 instances. No instance had all substances of an MDR defining class missing.
<sup>2</sup> All samples taken during and after the year 2020 were considered as after the start of the COVID-19 pandemic, while samples up to and including 2019 were considered before the start of the pandemic.
<sup>3</sup> Using logistic regression adjusting for age, sex and sample from ICU, taking multiple instances per patient into account.
Abbreviations: OR = odds ratio; 95% CI = 95% confidence interval; ICU = intensive care unit; N/A = not available.

In relation to this, the odds of resistance to tobramycin showed a significant reduction after the start of the pandemic (OR 0.24, 95% CI 0.09–0.66, p < 0.01), while resistance to amikacin also declined (OR 0.07, 95% CI 0.01–0.57, p = 0.01). For other agents, including cefepime, ceftazidime, ciprofloxacin, imipenem, levofloxacin, meropenem, and piperacillin-tazobactam, we did not find clear evidence of a difference in resistance after the start of the pandemic (**Figure 4**).

In none of the 492 instances were all resistance measures in one class missing, therefore leading to no missing data for MDR classification. Some missing data was observed for ceftazidime (1 instance) tobramycin (1 instance), levofloxacin (2 instances) and amikacin (247 instances). The high proportion of missing data for amikacin reflects its selective use, as it was not routinely applied to test aminoglycoside resistance.

### Antimicrobial consumption before and after the pandemic

Antibiotic consumption, measured in median days of therapy (DOT) per month to reflect the risk of development and selection of antimicrobial resistance as a function of exposure time, was compared before and after the start of the COVID-19 pandemic. Because of limited availability of historical data, the years 2018 and 2019 were classified as before pandemic, while 2020 to 2023 were considered as after the start of the pandemic. Antibiotics with an overall substantial decline in consumption after the start of the pandemic included amikacin, ciprofloxacin, gentamicin, levofloxacin, tobramycin, and piperacillin/tazobactam. No substantial change was observed for cefepime, ceftazidime, moxifloxacin, or meropenem (**Table 4** and **Figure 5 Panel A**).

**Figure 5.**
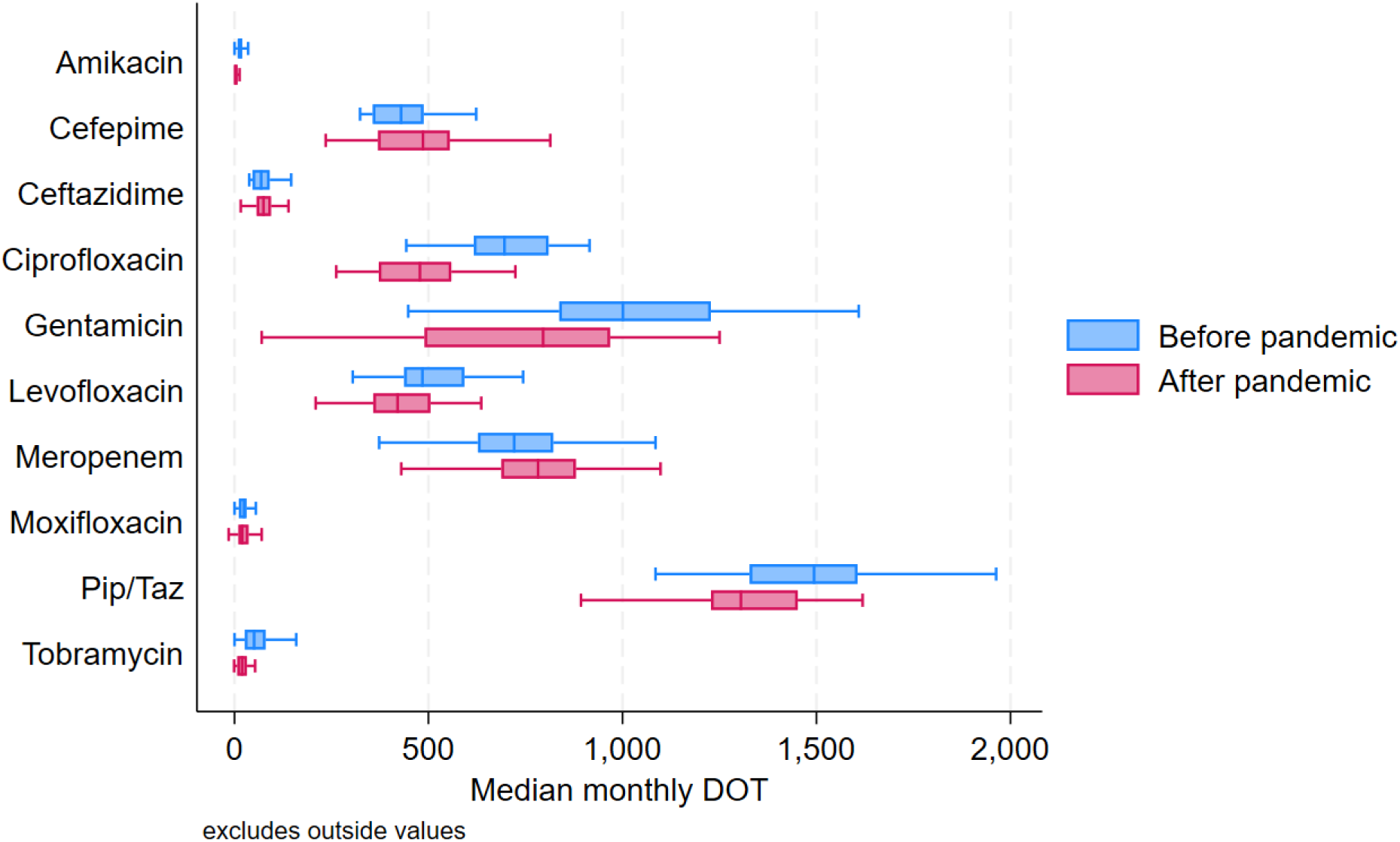
Panel A: Overall median antimicrobial days of therapy per month before and after the start of the pandemic. Box plots showing the median monthly antimicrobial days of therapy (DOT) for each substance before and after the start of the COVID-19 pandemic. Abbreviation: Pip/Taz = piperacillin-tazobactam

**Figure 5.**
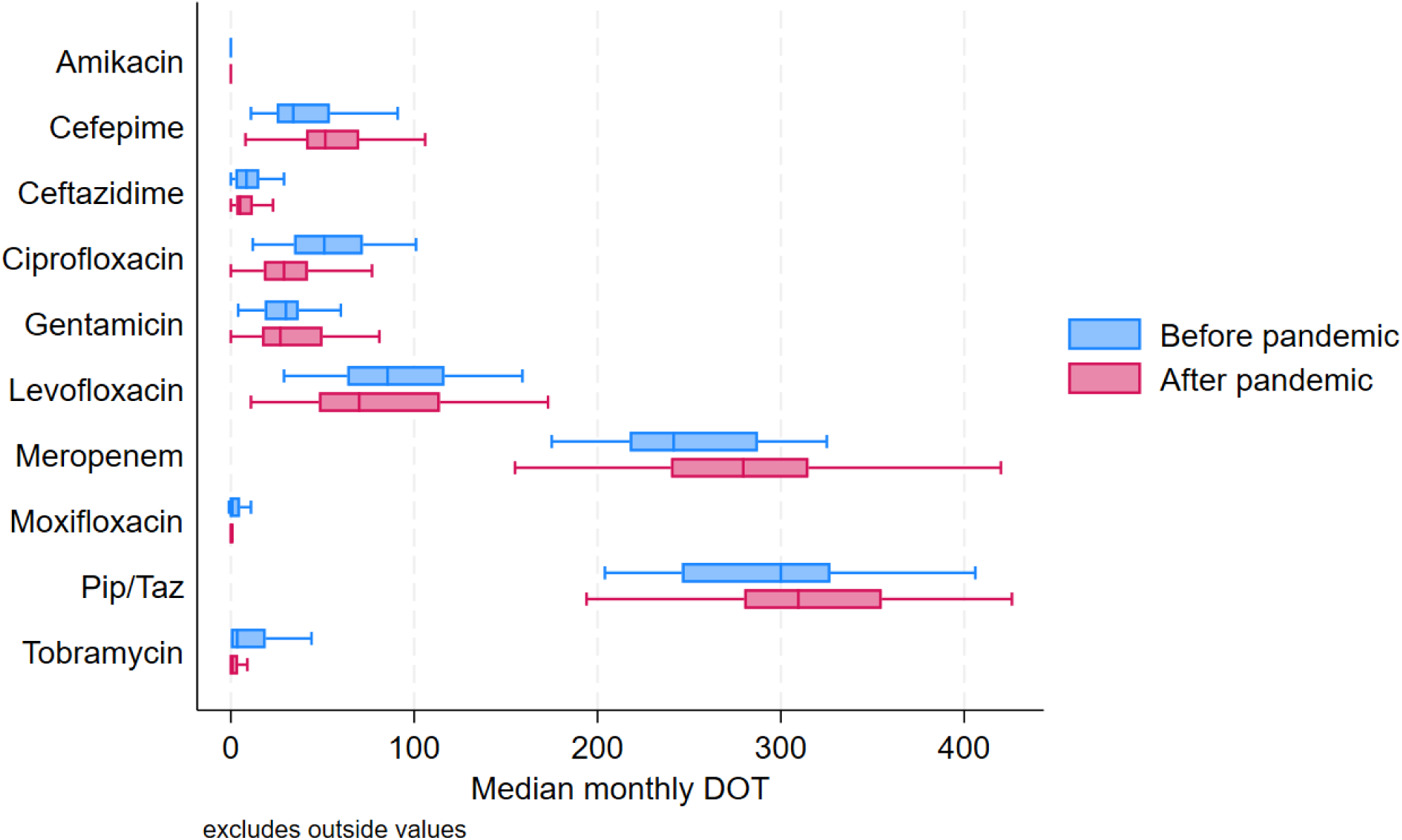
Panel B: Median antimicrobial days of therapy per month before and after the start of the pandemic on the ICU only. Box plots showing the median monthly antimicrobial days of therapy (DOT) for each substance before and after the start of the COVID-19 pandemic on the intensive care unit (ICU). Abbreviation: Pip/Taz = piperacillin-tazobactam, ICU = intensive care unit

**Table 4:**
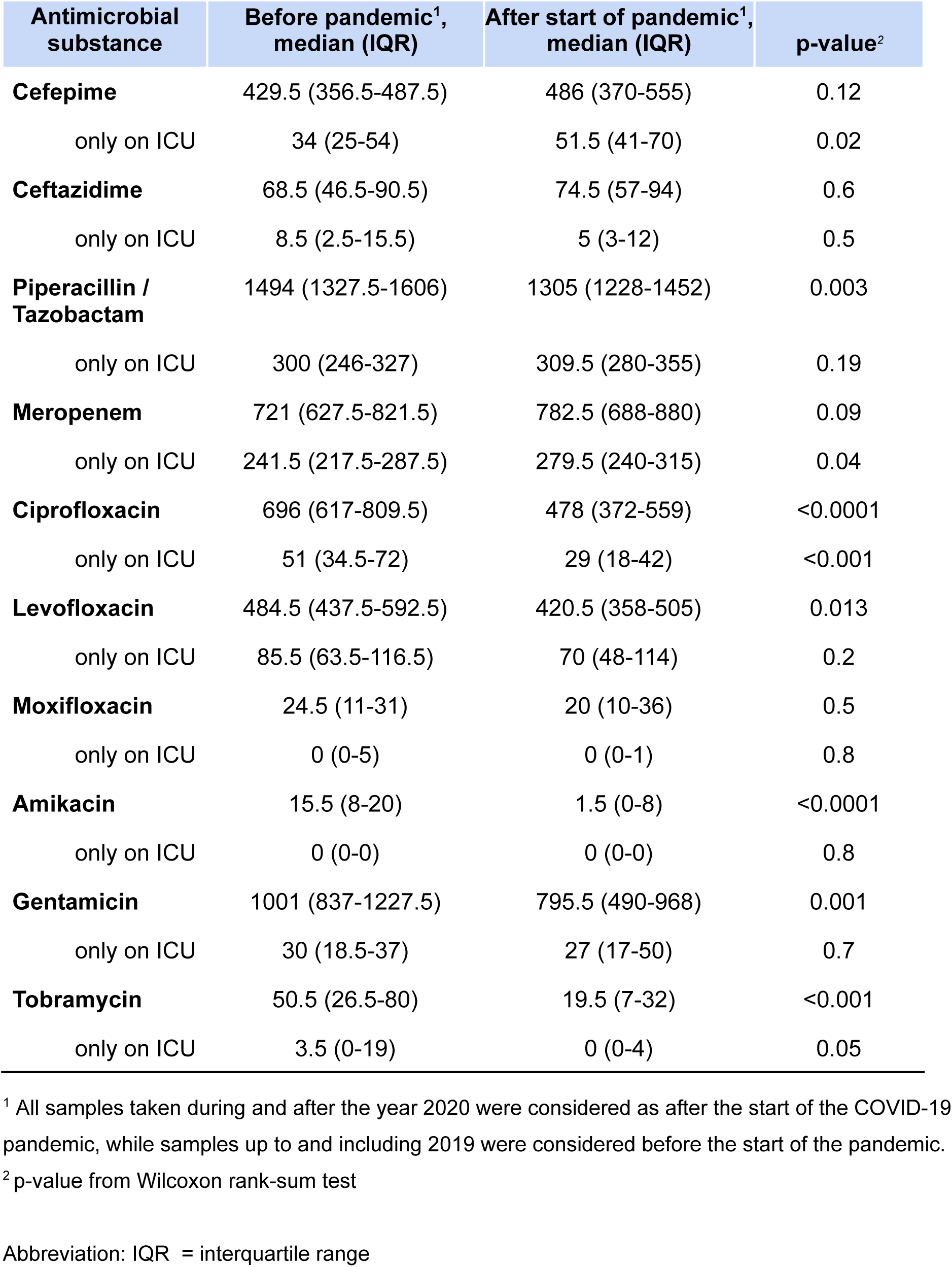
Median monthly antimicrobial days of therapy (DOT) before and after the start of the pandemic.

In the ICU, median monthly DOT increased for cefepime and for meropenem. In contrast, consumption decreased for ciprofloxacin, and for levofloxacin, when comparing the periods before and after the start of the COVID-19 pandemic (**Table 4** and **Figure 5 Panel B**). Consumption of ceftazidime, piperacillin/tazobactam, moxifloxacin, amikacin, gentamicin, and tobramycin remained similar.

## DISCUSSION

In this retrospective cohort study of 333 adult inpatients with PA bacteraemia at a tertiary university hospital over a 10-year period, we found an association between the time period after the the start of the COVID-19 pandemic and a decline in the occurrence of MDRPA isolates. We also observed substantial reductions in resistance to aminoglycoside antimicrobials, specifically tobramycin, and no significant changes in mortality among patients with MDRPA.

In alignment with our results, an Italian retrospective cohort study also reported a decrease in multidrug-resistant ESKAPEEc isolates, including PA, during the COVID-19 pandemic in a COVID-19-free ICU setting [14]. This suggests that heightened infection control measures and antimicrobial stewardship efforts during the pandemic likely contributed to the observed reductions in MDR. Similarly, a significant decline in invasive group A *Streptococcus* necrotising soft tissue infections was reported during the pandemic restrictions in our hospital, likely due to reduced transmission facilitated by isolation measures [15]. Together, these findings highlight the broader impact of pandemic-related infection control measures.

Moreover, shifts in patient populations and treatment priorities that began after the start of the pandemic appear to have influenced the epidemiology of nosocomial infections in our study. Notably, among patients with MDRPA we observed a lack of pulmonary comorbidities or cardiovascular diseases and reduction in burn injury and organ transplantation. The reduction in patients with chronic pulmonary conditions and cardiovascular diseases might partly reflect the high mortality rates among this vulnerable group during the pandemic. Among patients with organ transplantation, 5 out of 8 underwent a lung transplantation because of cystic fibrosis. All 5 patients experienced MDRPA bacteremia before the start of the pandemic. These changes persisted beyond the pandemic period, indicating a longer-term impact on hospital admission patterns. The rise in the proportion of malignancy cases, on the other hand, may reflect advances in oncology care in recent years, rather than being directly associated with the pandemic. Together, these findings highlight the complex and sustained effects of both pandemic-related and independent factors on patient populations and nosocomial infection risks.

Our study’s findings also provide important context for understanding MDRPA in a global setting. A recent retrospective cohort study in children from Israel reported that MDRPA accounted for 8% of all *Pseudomonas* isolates, compared to 12% in our adult cohort [16]. Despite differing patient populations, both studies highlight the ongoing clinical challenge posed by MDRPA, particularly in terms of extended hospital stays and the high proportion of ICU admissions among affected patients.

The decline in MDRPA after the start of the pandemic occurred alongside shifts in antibiotic consumption. In our cohort, consumption of aminoglycosides, fluoroquinolones and piperacillin/tazobactam decreased substantially, likely contributing to these reduced resistance rates. Conversely, the increased use of cefepime and meropenem in the ICU may be due to suspected pulmonary superinfections in COVID-19 patients, underscoring the complexity of antimicrobial stewardship efforts during the pandemic and the importance of sustained post-pandemic stewardship programs.

The reduction in certain antibiotics observed in our study aligns with broader global trends of decreased antibiotic consumption during the pandemic, particularly in high-income settings [17]. However, while also a rebound in consumption has been reported [17], our data indicate that reductions in specific antibiotics, like tobramycin and ciprofloxacin, persisted post-pandemic, potentially contributing to sustained lower rates of MDRPA. Especially the decline in ciprofloxacin use is relevant, as it remains the only oral treatment option for PA bacteraemia.

Our study has several strengths. First, the 10-year timeframe allowed for robust temporal comparisons, capturing the dynamics both before and after the start of the COVID-19 pandemic. Second, our use of a comprehensive electronic medical records database ensured high-quality data collection. Third, including all instances of PA bacteraemia and accounting for clustering of instances within patients in our primary analysis minimised bias from instance selection in evaluating the association between pandemic-related factors and MDRPA occurrence. Fourth, the availability of patient-level antimicrobial prescription data allowed us to directly evaluate temporal selection pressure using days of treatment (DOT) as the measure of antimicrobial consumption. Finally, our sensitivity analysis confirmed that the change in MDRPA classification criteria did not influence the study’s conclusions.

Nonetheless, our study also has limitations. First, the retrospective design precludes causal inference, and unmeasured confounding, such as prior antimicrobial exposure or colonisation with MDR pathogens. Second, other important differences in unmeasured patient characteristics could have influenced the observed associations. Third, we lacked data on the total number of patients treated in our hospital, therefore only allowing us to measure the occurrence as proportion of MDR in PA bacteremia, preventing us from drawing conclusions about incidence. Fourth, we only had clinical data on patients with MDRPA bacteremia and not all patients with PA bacteremia, making it impossible to identify any changes of risk factors contributing to the occurrence of MDR. Fifth, we did not have data on total patient days, preventing us from calculating the density of antibiotic use as DOT per 1000 patient days. However, while the hospital’s capacity remained relatively stable throughout the study period, patient discharges decreased slightly from 42,376 in 2018 to 39,153 in 2023 [18]. This modest decline in discharges is unlikely to explain the observed reduction in antibiotic consumption, minimising potential bias from changes in total patient days. Sixth, we were not able to measure changes in patient behaviour after the start of the pandemic, which might also have affected antimicrobial consumption and/or resistance selection. Finally, while we observed significant reductions in resistance to certain antibiotics, the impact of these changes on clinical outcomes, such as mortality or treatment success, cannot be deduced from our study.

## CONCLUSION

In conclusion, we found evidence for a decline in the occurrence of MDRPA bacteraemia in association with the time period after the start of the COVID-19 pandemic. These findings suggest that pandemic-related changes in infection control, antimicrobial use and population demography may have contributed to improved resistance profiles in *P. aeruginosa* bacteremia. Further research is needed to confirm these trends in other settings and to explore the long-term impact of sustained post-pandemic interventions on AMR. Nevertheless, our findings suggest that alongside changes of antimicrobial consumption as seen with antimicrobial stewardship programmes and rigorous hygiene measures could reduce MDRPA in bacteraemia within a relatively short time frame.

## Data Availability

The datasets used and/or analysed during the current study are available from the corresponding author upon reasonable request.

## ABBREVIATIONS

MDRPA: Multidrug-Resistant *Pseudomonas aeruginosa*
PA: *Pseudomonas aeruginosa*
WHO: World Health Organization
CDC: Centers for Disease Control and Prevention
SARS-CoV-2: Severe Acute Respiratory Syndrome Coronavirus 2
ICU: Intensive Care Unit
HR: Hazard Ratio
CI: Confidence Interval
OR: Odds Ratio
IQR: Interquartile Range

## DECLARATIONS

### Ethics Approval

The ethics committee of the Canton Zurich reviewed the study protocol (Kantonale Ethikkommission Zurich BASEC ID Req-2022-00076) and concluded that the study did not meet the criteria for requiring ethical approval within its remit. All research was carried out in accordance with Good Clinical Practice (GCP) standards. The data required for this study were anonymized before their use.

### Consent for publication

Not applicable.

### Authors’ contributions

Conceptualization: YB, PMF and SDB. Funding acquisition and resources: SDB. SDB was responsible for the ethical approval. PMF performed the statistical analysis, while LP contributed to plotting figures. YB and MP collected the data. YB, PMF, SM and SDB interpreted the data. YB, PMF and SDB wrote the first draft of the manuscript. All authors critically reviewed the manuscript and approved the final version.

### Tools for Analysis and Manuscript Preparation

All analyses were performed using Stata 18 (Stata Corporation, College Station, TX, USA), while Figure 4 was created using R version 4.2.1 with the ggplot2 package [19]. During the preparation of the manuscript, ChatGPT (GPT-4o) by OpenAI, and Google Documents were employed for text editing and drafting purposes. ChatGPT aided in enhancing the language clarity in certain paragraphs and improving the text drafting process, while Google Documents aided in real-time collaboration and efficient document management. It is important to note that ChatGPT was not used for analysis, intellectual content or literature review. The authors have thoroughly reviewed and verified any passages edited by ChatGPT, taking full responsibility for the manuscript’s overall quality and accuracy.

## Acknowledgments

We would like to thank Christopher Witzany for his critical review of this manuscript and helpful comments to further improve the quality of our work.

